# Development and Validation of a Culturally Tailored Mental Health Literacy Training Program for Indian School Teachers

**DOI:** 10.64898/2026.09.15.26363096

**Authors:** Regina Shoba Dass, P Chinnadurai

## Abstract

**Background:** School teachers are the first responders in the early detection and referral of students with mental health issues and there is an urgent need for Mental Health Literacy training to support this. Childhood and adolescent mental health have emerged as a national health problem, so there is a need to plan an intervention program on mental health literacy for teachers in the schools of India as there are very few planned interventions.

**Objective:** To design and test an intervention package for school teachers regarding mental health literacy.

**Materials and Methods:** The training module on mental health literacy for school teachers was developed through a needs assessment, by conducting in-depth interviews with school teachers (n = 12), gathering experts’ opinions on the content for the training manual (n = 12), and content validation of the training manual by experts (n = 7). Content validation index and kappa value was used for validation of the manual.

**Results:** School teachers were targeted in the development of the components of the training manual to orient them to mental health and mental illness, and to prepare them to manage school children with behavioural and mental health disorders and know when to refer. The contents covered the introduction to mental health and mental disorders, myths and stigma related to mental illness, common developmental, behavioural and mental health disorders in children, role of teachers, mental health first aid, and seeking help and finding support. The Kappa value was modified (K = 0.932) and showed that the content was very relevant to use for school teachers.

**Conclusion:** School teachers’ mental health literacy programs in India are vital interventions to address the mental health crisis in the country. With the development of structured MHL modules, India can start to overcome this huge treatment gap by enabling educators to become effective early intervention points.

## Introduction

Mental health disorders are a significant burden in the world, with about 50% of all cases appearing before the age of 14 and about 75% by the age of 25 (Kutcher & Wei, 2020; Patel et al., 2018). The developmental stages are especially important because they represent formative periods that can have a profound impact on psychological health and wellbeing in the long term (Ilango et al., 2025; Roy et al., 2019). Nearly 50 million children in India are suffering from mental health problems, with 23.33% of them in school settings and as high as 13.5% among the urban population (Ilango et al., 2025; Kamath et al 2021; Parikh et al., 2019). There is a very high demand and supply gap for children with mental illness globally, with only a small proportion of children who need care receiving professional mental health support (Venkataraman et al., 2019) and in India, a significant proportion of children have mental illness, but do not receive any mental health services (Mastrorio et al., 2020; Parikh et al., 2019). This deficit is exacerbated by the lack of trained professionals, as in India, there are 0.4 psychiatrists per 100,000 people (Siraj et al., 2024; Vieira et al., 2014).

Schools are also key players in mental health promotion and prevention (Kutcher & Wei, 2020; Whitley & Gooderham, 2016), as they spend a large part of their childhood years in school. Teachers, who are at the frontline of the classroom, can easily observe subtle changes in students’ behavior, mood, social interactions and academic performance that are indicative of emerging distress (Giri et al., 2025; Gunawardena et al., 2024). However, many teachers report being overwhelmed and confused by the differentiation process when trying to distinguish between normal adolescent changes and clinical manifestations of mental disorders (Giles-Kaye et al., 2022; Levkovich & Hazan-Liran, 2026; Shelemy et al., 2019). Most teachers are inadequately prepared for mental health issues, with most not receiving sufficient pre-service training in mental health (Kutcher & Wei, 2020; Mastrorio et al., 2020; Whitley & Gooderham, 2016).

As long as teachers do not have a structured mental health literacy, they are more likely to use office disciplinary referrals as a method of identifying a mental health problem that will miss signs of more internalizing issues, such as anxiety, depression, and other disorders (August et al., 2017; Baxter et al., 2022; Jose et al., 2025). The modern generation gap and differential exposure to digital media also heighten the challenges that educators face in identifying students’ emerging distress (Cooper et al., 2025; Duraku et al., 2023). Additionally, having low mental health literacy is directly associated with elevated stigmatizing attitudes that have significant impacts on silencing youth and reducing their ability to seek help (Baxter et al., 2022; Kashyap et al., 2024).

In the Indian context, culturally tailored interventions hold special significance because the widely-used generic models from high-income countries do not adequately meet the need for addressing the pervasive stigma, classroom diversity, an exam-oriented academic pressure and unique family structures (Harte & Barry, 2024; Jose et al., 2025; Kågström et al., 2023; Kaur et al., 2023). The most effective teacher training should go beyond focusing on knowledge and theory, and include practical, skills-based strategies and the creation of formal pathways between schools and community mental health services (Afshari et al., 2022; Wei et al., 2011). The sustainable implementation involves national initiatives like Rashtriya Kishor Swasthya Karyakram, along with school and mental health provider collaboration (Margaretha et al., 2023; Parikh et al., 2019; Roy et al., 2019).

This study attempts to create a culturally adapted trainable program in mental health literacy for school teachers (STs) in India with the view of bridging gaps between awareness and appropriate referrals for treatment in schools, based on the expert recommendations, with the intent to use schools as life-saving intervention centres for adolescent well-being.

## Materials and methods

The Institute Ethics Committee approved the study (Ref No□CMRU/SLS/RGS/2024/22CPHDF027). From 2024-2026 as a part of a PhD study the development of a training program for Mental Health Literacy for school teachers was conducted. The training module is clearly targeted at teachers in state and CBSE board schools in Karnataka. A total of 12 school teachers (n =12) were chosen to participate in an in-depth interview by the first Author to determine the training needs of the school teachers with regards to Mental Health Literacy. Themes were generated from the interview using thematic analysis, which resulted in seven Primary themes. In addition, 16 professionals from the psychiatry, psychology, psychiatric social work and educational areas were approached in order to gain their input for the content of the training programme, and six themes were derived from the qualitative analysis of the interviews with the key informants. Training program contents were derived from the need assessment of school teachers and the expert opinion which included introduction to mental health and illness, myths and stigma associated with mental illness, Psychological First Aid, seeking help, identification of common problems in children and their management - developmental disorders, common mental disorders, school related problems.

Content validation of the training program was done by following six steps as shown in Figure 1, preparation of the form for content validation, selection of the content experts, content validation, review and scoring of each item, and calculation of item-level content validation index (I-CVI = the number of experts who agree / the number of experts). Seven experts gave consent to validated the manual. The accepted CVI is ≥0.83, when six to eight experts validate the content, (Lynn MR., 1986). The content of the training program was designed based on need assessment and expert opinion and presented to the experts for review. The following rating scale was used to rate each of the training modules by each subject expert: ‘not relevant’, ‘somewhat relevant’, ‘quite relevant’, ‘very relevant’. The content analysis was conducted with the experts’ ratings. The suggestions and recommendations of the experts were taken on board.

Content validation was performed by seven subject experts (two psychiatrists, one clinical psychologist, one psychiatric social work faculty member, one school counsellor and two school principals). The number of years the experts had ranged from 5 to 15 years. The intervention package was content validated using expert feedback with the ABC Content

Validation Index (CVI) for item-level relevance, suitability and appropriateness. This was computed as item-level CVI (I-CVI) that is the percentage of experts who checked “relevant” in the corresponding column. Items with an I-CVI of > 0.79 are considered relevant, I-CVI range of 0.70-0.79 means the item requires revisions and I-CVI < 0.70 means the item should be eliminated. Average and universal agreement were used to obtain scale-level CVI. To account for chance agreement, modified Kappa (κ) was calculated for each item; values of κ > 0.74 were considered excellent, κ = 0.60–0.74 good, and κ = 0.40–0.59 fair. This two-stage process provided quantitative rigor and the ability to incorporate the suggestions of experts in refining the package (Polit, D. F., & Beck, C. T., 2006). The training program has five modules and seven sessions, with each session taking 45 minutes to 1 hour. Informed written consent was obtained and a pilot testing of the training module was carried out with 10 school teachers. Feedback was obtained from the participants after each session.

## Results

To assess the need for training teachers in Mental Health Literacy, an in-depth interview was conducted with twelve school teachers. Seven themes emerged through thematic analysis. The themes included conceptualizing mental health in students, psychological problems among students, contributing factors to children’s behavioral and emotional problems, strategies for managing behavioral and emotional problems, challenges in managing behavioral and emotional problems, need for capacity building in addressing problems, and varied exposure and understanding of mental disorders.

Sixteen subject-matter experts with five or more years of experience in Psychiatry, Psychology, and education were interviewed to provide their opinions on the content to be used in developing modules for a training program for teachers on MHL. Thematic analysis of the expert opinions yielded six themes: common problems in help-seeking, teacher-reported problems, key challenges faced by teachers, effective strategies for supporting teachers, core areas for teacher training, and extended suggestions for teacher.

**Figure 1.**
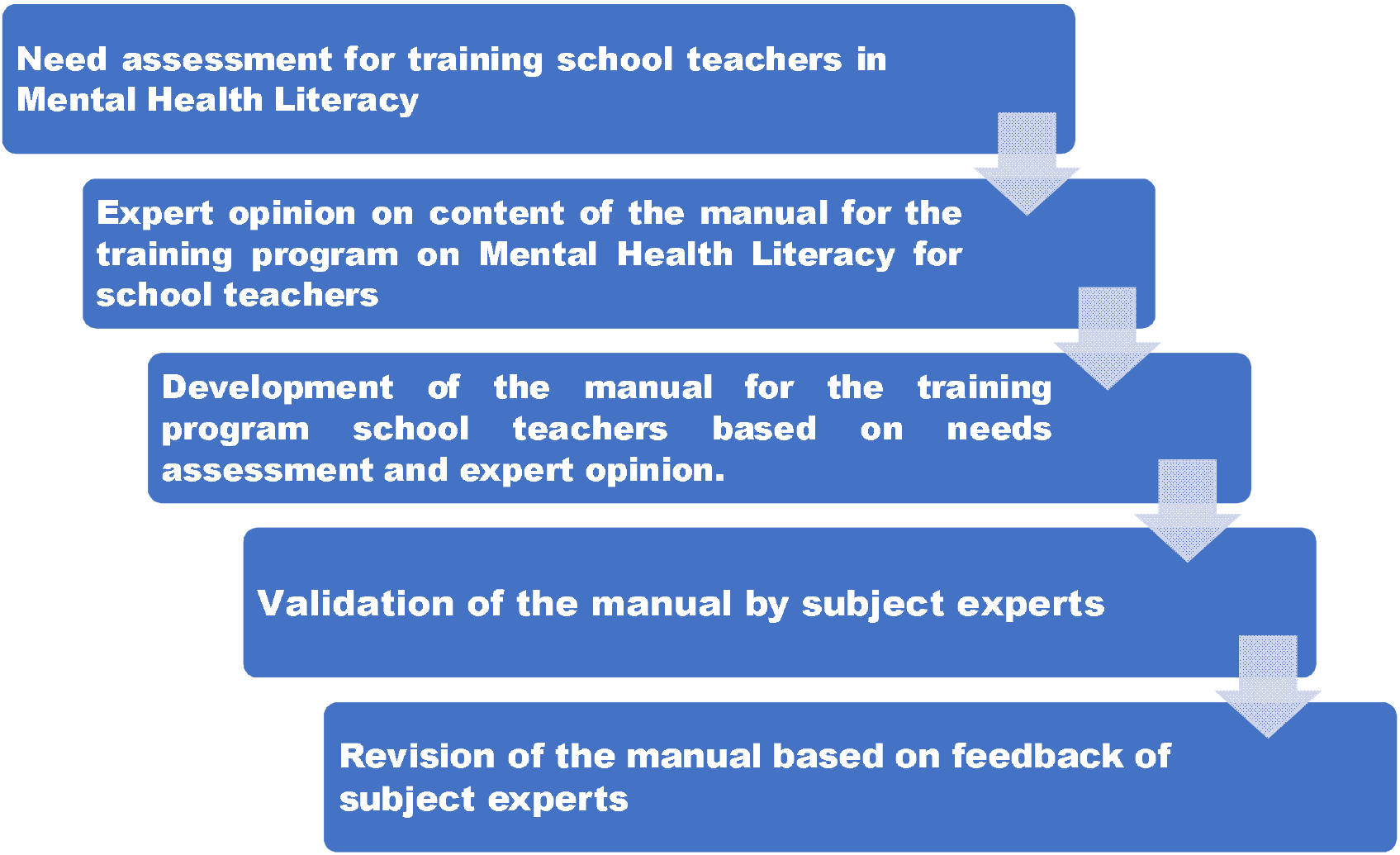
Development Process of the Training Manual.

### Development Process of the Training Manual

All experts accepted that the modules on Introduction to mental health and mental illness, Mental Health First AID, seeking help & finding support, and common childhood problems (developmental, behavioural, and mental disorders) and their management, were relevant.

Five experts reported that the module on myths and stigma related to mental illness, was quite right. Overall, there was a general conscience regarding appropriateness of the modules.

Content of group intervention sessions Table 3 reveals the index of content validity (CVI) and kappa score of the group intervention manual. The results of the inter-rater reliability among seven experts for the relevance of the modules on Mental Health Literacy Training Program for School Teachers is shown in Table. The agreement beyond chance was not statistically significant with overall Kappa coefficient of −0.0343, Z-value of −0.42 and a p-value of 0.661.

**Table 1.**
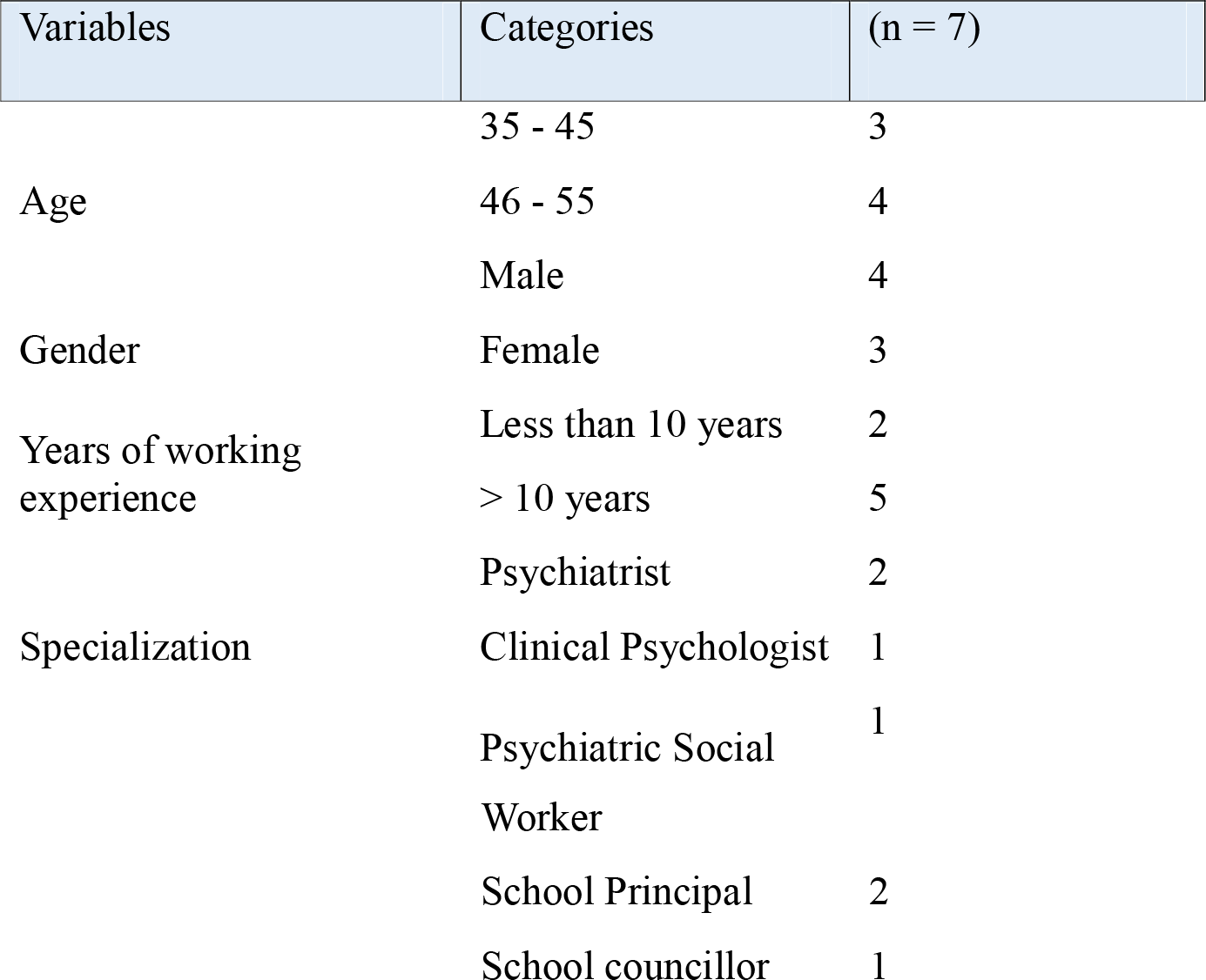
Profile of subject experts. Table 1 shows the profiles of the subject experts who participated in the content validation of the group intervention manual. Five experts who validated the module were aged from 30 to 40 years old, and four were males. Five experts had more than five years of work experience in mental health and intimate partner violence. Four experts were teaching and practicing in a mental health institute.

**Table 2.**
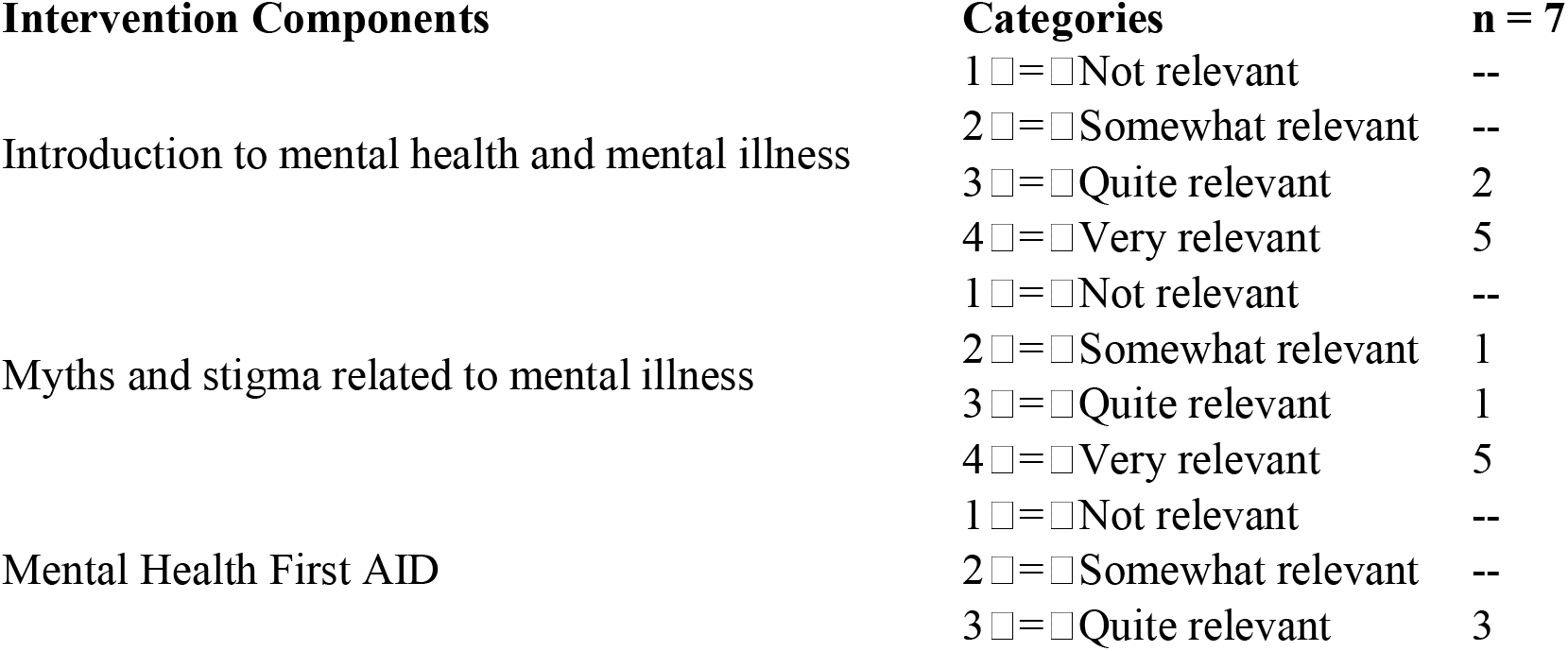
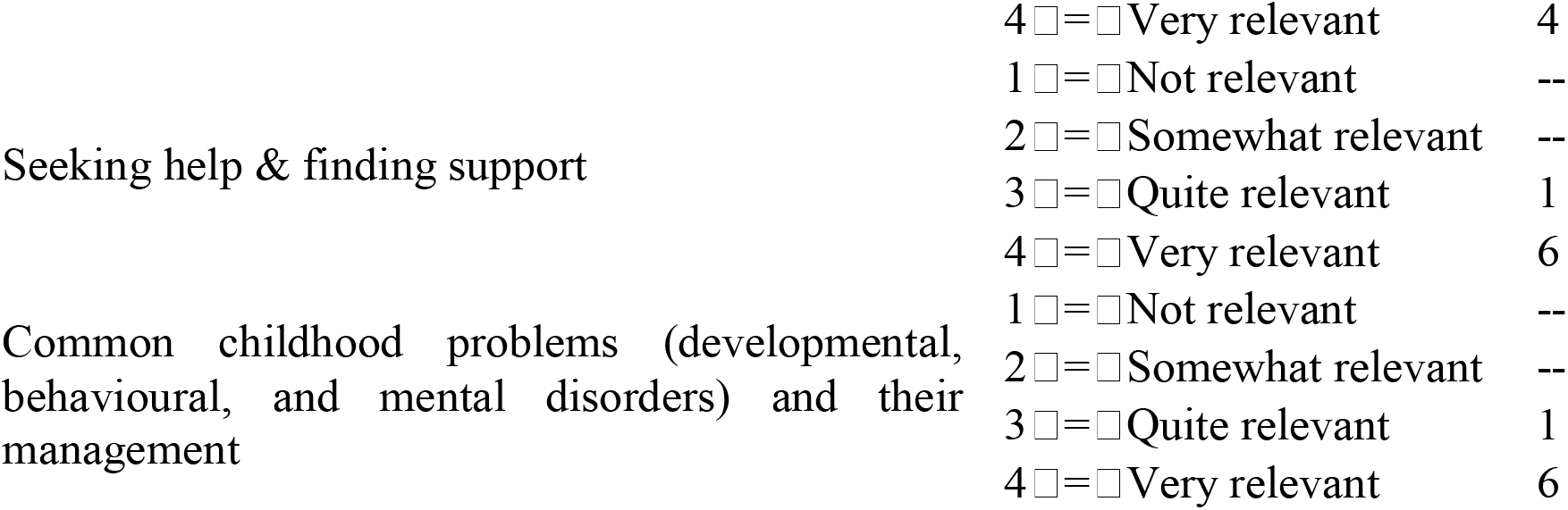
Content validation of the Training Program.

The range of the category-specific Kappa coefficients was between −0.1048 and 0.1162. The highest agreement was seen in the category Somewhat Relevant (Rating = 2), with a Kappa coefficient of 0.1162, the lowest agreement was seen in the Very Relevant (Rating = 4) with a negative Kappa coefficient (κ = −0.1048). There were no statistically significant differences (p > 0.05) found between the category-specific agreements.

Limited differences among experts’ ratings and the relatively low number of items rated could be the reason for the low Kappa values. However, in content validation studies, experts may give very close ratings, give high Kappa values, but observe agreement is not that high. Thus, the Content Validity Index (CVI) and Modified Kappa (K*) are usually more suitable indicators of content validity.

The overall inter-rater agreement of the seven experts on the relevance of modules on Mental Health Literacy Training Program was not statistically significant (κ = −0.0343, Z = −0.42, p = 0.661). Content validity, however, of the module was excellent as evidenced by Scale-Level Content Validity Index (S-CVI/Ave = 0.943) and Modified Kappa (K* = 0.932) which meant that the content was very relevant and suitable for use by teachers in schools.

### The content of the training program manual

#### MODULE I-SESSION 1: Introduction to mental health and mental illness

The aim of this session is to provide teachers with a clear understanding of mental health and mental illness. Teachers will be introduced to the characteristics of a mentally healthy person, the causes and types of mental illness, and available treatments. The session also aims to help teachers recognize common mental health problems and their symptoms in students. Small group discussions will focus on the impact of mental health on students, and participants will learn about the components of health, characteristics of mental well-being, causes and classifications of mental disorders, and general treatment approaches. This session lays the groundwork for future sessions.

#### MODULE II-SESSION 2: Myth and stigma towards mental illness

The aim of this session is to dispel myths and reduce stigma related to mental illness among teachers. It seeks to foster an evidence-based understanding and equip teachers with strategies to combat stigma in the school environment. Group discussions covered myths about causes and treatment of mental illness, stigma, ways to overcome it, and teachers’ roles in reducing student stigma. Stigma is a primary barrier to youth seeking care, so addressing teachers’ misconceptions benefits students. The session concluded by preparing teachers for the next module on child disorders.

#### MODULE III: Developmental, behavioral & mental disorders in children and their management. This module builds on earlier sessions to introduce specific challenges related to children. SESSION 3: Developmental disorders

The aim of this session is to equip school staff with knowledge about common emotional, behavioral, and mental disorders in children and with practical classroom strategies for addressing them. Teachers will learn to identify these disorders, manage classroom issues, and know when to refer students for further help. Discussion and role play will center on intellectual disabilities, autism spectrum disorders, learning disabilities, and speech and language disorders.

#### MODULE III-SESSION 4: Classroom strategies to manage common emotional and mental disorders in children

The aim of this session is to provide teachers with classroom management strategies to address common emotional and behavioral disorders such as anxiety, separation anxiety, depression, ADHD, and conduct problems. Teachers will participate in case studies to apply these strategies, recognize behavioral indicators, and determine when to refer students for further support.

#### MODULE III-SESSION 5: School related problems and their management

The aim of this session is to prepare teachers to identify, respond to, and support students facing bullying, substance abuse, media and internet addiction, cyberbullying, abuse, or trauma. Teachers will practice discussing and handling these issues, including working with parents, through scenarios and role-play.

#### MODULE IV-SESSION 6: Mental health first aid (psychological first aid)

The aim of this session is to provide teachers with practical skills and steps for offering psychological first aid to students experiencing acute emotional or mental health issues. Teachers will learn how to listen non-judgmentally, assess risk, reassure students, encourage self- and professional help, follow up, and support ongoing treatment. The session will also address collaboration with families and caregivers.

#### MODULE V-SESSION 7: Help-seeking and finding support

The aim of this session is to enable teachers to recognize when and how to seek help and support for students, themselves or others for mental health concerns. This session aims to help teachers understand that stressful life events may require seeking help, to inform them whom they can approach if they have concerns about themselves or others, to outline school-based mental health support personnel, and to familiarize them with available community resources and helplines. This session integrates previous learning and emphasizes available support pathways.

## Discussion

This study highlights an important paradox: teachers are known to be the first point of contact for students’ mental health care, yet do not typically have the capacity to provide specialized mental health services in schools (Gunawardena et al., 2024; Shelemy et al., 2019). The thematic analysis of the expert interviews revealed six main themes which directly informed the design of the training program: Common problems in help-seeking, Teacher-reported problems, Key challenges faced by teachers, Effective strategies for supporting Teachers, Core areas for Teacher Training, and Extended Suggestions for Teacher. Themes converged to a consensus that providing these teachers with effective change agent training requires interactive, robust training sessions with practical information on early intervention and not adding the load of being a therapist to their teaching responsibilities (Karim, 2024).

One very common theme in the discussions was the need for more training and understanding and the realities of staffing shortages in the school mental health system (Giles-Kaye et al., 2022; Gunawardena et al., 2024). Additionally, the use of knowledge transfer was identified as an ineffectiveness to resolve the stigma associated with seeking help and the increasing generation gap between teachers and students who are immersed in digital technologies (Esteban et al., 2025; Cooper et al., 2025; Manjubairavi et al., 2025). These barriers exacerbated the risk of teaching staff using informal (and potentially unreliable) identification systems, including office disciplinary referrals, which were more likely to capture externalising behaviours than internalising distress (Baxter et al., 2022; Jose et al., 2025).

As a community, the experts identified a number of aspects as being critical for a culturally responsive module. They included an understanding of normal growth and development, awareness of some behavioural, emotional and mental health issues in children, practical classroom management skills, referral skills, digital empowerment and collaboration with parents and community, and explicit attention to teacher well-being. This composition aligns with the existing frameworks, such as Kutcher and Wei’s (2020) three critical core elements of school mental health, which include mental health literacy for teachers, pre-service and in-service mental health teacher training, and integrated school-site provision of mental health care.

Our results corroborate findings from other countries in the world; recent meta-analyses have indeed shown that mental health literacy interventions can indeed empower teachers to promote students’ well-being, but with varying degrees of sustainability. Some studies have also reported small but significant immediate impacts on teachers’ mental health knowledge and reducing stigma (Liang et al., 2026; Liao et al., 2023). Importantly, however, although the effects of knowledge improvements are generally stable over time, the effects of attitudinal changes can be less consistent, suggesting that single-shot interventions may not be enough to sustain a change in attitude over time (Liang et al., 2026; Liao et al., 2023). In addition, there exists a close relationship between MHL training and helping behaviors – the practical action that teachers take to support students – and the helping behaviors have a significant improvement after teacher structured training (Liang et al., 2026).

Recent trials in the Indian context are quite supportive of an empirical basis. After a modular MHL trainings among Indian high school teachers, significant increase in knowledge (mean score change of 3.34) and attitudes and beliefs (increase of 0.95) (Prabhu et al., 2025). In Varanasi, body language, teacher-student relationship and the application of effective teaching methods in classrooms, through sensitization programs, was seen to have a great impact on teachers in government schools to support well-being (Siraj et al., 2024). The “Teachers Leading the Frontlines” model in Darjeeling achieved reduction in mental health symptom severity (Cohen’s d = 0.70; p = 0.024), higher academic performance in math (d = 0.63; p = 0.0006), and higher academic performance in reading (d = 0.83; p = 0.001) among students (Giri et al., 2025). The converging results indicate that evidence-based models can be transferred to various cultural and economic contexts (Liao et al., 2023) as long as they are adapted to the local linguistic, cultural and systemic realities (Harte & Barry, 2024; Kågström et al., 2023).

The key takeaway from these comparative results is that the need to shift from isolated use of modules to a whole school approach (Margaretha et al., 2023; Patel et al., 2018). Formal partnerships between schools and community mental health providers are needed to implement such an approach in accordance with the “School Based Pathway to Care” model, which focuses on identification and linkage, with teachers not providing treatment (Wei et al., 2011). Child psychiatrists or clinical psychologists are also scarce in low-resource contexts such as India, and these partnerships are especially relevant given the need to ensure sustained referral pathways in such settings where there is limited capacity (Margaretha et al., 2023; Roy et al., 2019). Incorporating MHL with pre-service training (such as D.Ed., B.Ed. and B.P.Ed.) is also crucial, enabling future teachers to identify and address mental health issues (Mastrorio et al., 2020; Whitley & Gooderham, 2016).

Educational institutions can create proactive environments where help-seeking is normalized by placing sustainable frameworks and mental health first, not last, in their school culture, rather than as an afterthought (Cooper et al., 2025; Lakind et al., 2023). This systemic shift is based on task-shifting approaches that address gaps in resources and defines the role of the teacher in early detection while distributing specialized support to trained professionals (Duraku et al., 2023). In the end, these methods could best be understood as a multi□layered support system for schools to consistently engage at□risk youth in early intervention (August et al., 2017; Childs□Fegredo et al., 2020).

### Recommendations

For Research the MHL training module needs testing with larger and more diverse numbers of teachers from both government and private schools, rural and urban schools, and to enhance statistical power and generalizability. To make causal inferences and to reduce selection bias, randomized controlled trials are required. Follow-up assessments at six and twelve months after the intervention could help determine if the improvements in knowledge, stigma reduction and helping behaviors were maintained over time. Replication across culture and language would further support the adaptability of the module to the context of education.

When training is integrated in school culture, as opposed to viewed as an isolated event, successful training implementation increases significantly (Baffsky et al., 2022; Patel et al., 2018). In addition to the group-based care techniques, a whole-school approach involving parents, school counselors, school administration, and community stakeholders should be adopted and parental engagement should be continued throughout the school year (Margaretha et al., 2023). Mental health champions (strategic leads responsible for implementation and effectiveness monitoring) offer the required organizational structure for sustainability (O’Reilly et al., 2018; Vay et al., 2024). In addition, teachers need to be trained on the ability to identify student needs and to connect identified students with formal health care, eliminating clinical responsibilities from teachers (Wei et al., 2011; Afshari et al., 2022).

Systemic integration (Mastrorio et al., 2020; Whitley & Gooderham, 2016) requires the embedding of MHL modules into pre-service and in-service teacher training programs (D.Ed./B.Ed. /M.Ed.), as well as in teacher training institutes. That identification efforts should translate into tangible pathways for evidence-based mental health care requires formal agreements between educational institutions and community-based mental health providers (Baxter et al., 2022; Margaretha et al., 2023). The implementation of the frameworks should consider the unique developmental vulnerabilities of adolescents, especially from upper primary schools to secondary and higher secondary schools (Ilango et al., 2025; Malar et al., 2025). When these interventions are linked to national initiatives like Rashtriya Kishor Swasthya Karyakram, they become integrated in the Indian public health discourse and can impact change over the long term (Parikh et al., 2019; Roy et al., 2019). Moreover, investing in technology-enabled platforms can enhance remote supervision and high-fidelity training on a large scale, particularly in rural or under-resourced areas (Frazier & Fosco, 2024; Hamdani et al., 2021).

### Limitations

#### Limited Generalizability

The module was used with a small number of teachers, which decreases the generalizability of the findings, again as would be expected in a pilot study. Future studies should be done using larger and more diverse samples to confirm these preliminary results (Liang et al., 2026; Liao et al., 2023).

#### No Randomization

Randomization was not used, limiting the ability to make causal inferences about the effects of the intervention on knowledge, attitudes, or helping behaviors. Further research needs to be conducted using cluster-randomized controlled designs to enhance causal inference (McGuckin & Leavey, 2021; Prabhu et al., 2025).

#### Short-Term Outcomes Only

This study will emphasize the immediate post-intervention outcomes. At this time, it is unknown whether any observed improvements in stigmatizing attitudes and helping behaviors are sustained over time without additional follow-up. The results indicated that knowledge gains are relatively stable; however, attitudinal and behavioral changes will need to be reinforced over time (Liang et al., 2026; Liao et al., 2023).

#### Self-Report Bias

Teacher self-reports also raise the possibility of social desirability bias (Giles-Kaye et al., 2022; Kashyap et al., 2024) which is common in self-report measures, as participants may overestimate their knowledge or attitudes to make them appear more positive than they actually are.

#### Selection Bias

These samples were purposive both for teachers and experts, and there is a possibility of selection bias as people who were willing to participate may already have positive attitudes toward MHL or be more involved in the conversation about mental health (Parikh et al., 2019).

### Contextual Limitations

#### Cultural and Linguistic Transferability

The module was written and trialed in a particular context of Indian school. Its key advantage is its foundation in Indian cultural schema, but the diversity within India (linguistic, regional, religious, and urban-rural) indicates the need to adapt it further to be replicated across different states (Harte & Barry, 2024; Kågström et al., 2023).

#### Limitations

The module covered only mental health literacy and not more complex and severe psychiatric illnesses, nor the systemic issues that teachers have to contend with in resource-stressed schools, including large class size, exam pressures, infrastructure issues, etc. (Harte & Barry, 2024; Jose et al., 2025; Mehra et al., 2022).

#### Stakeholder Scope

Only teachers were targeted. Parents, administrators and counselors are not systematically trained, leaving other components of the referral chain underdeveloped (O’Reilly et al., 2018; Parikh et al., 2019).

### Contextual Strengths

It is noteworthy to note that the cultural anchorage of the module is also its main strength. This module is also able to tackle issues of Indian stigma patterns, multilingual delivery, and integration with the existing pedagogical architecture, which is often missing from generic curricula (Harte & Barry, 2024; Jose et al., 2025; Kågström et al., 2023; Kaur et al., 2023).

## Conclusion

The purpose of this study was to develop and test a culturally appropriate mental health literacy program for Indian school teachers. Schools are especially well suited to act as first points of contact for children in need of mental health treatment, and there is a growing treatment gap in Indian child mental health (Giri et al., 2025; Margaretha et al., 2023; Roy et al., 2019), making the need for such a program timely. In a qualitative, phenomenological design, in-depth interviews were conducted with 16 multi-disciplinary experts to map the content required and content constraints of effective MHL training in Indian schools. The resulting module, consisting of 7 sessions, across 5 thematic areas (Introduction to Mental Health; Mental Health Myths and Stigma; Recognition of Emotional, Behavioral and Developmental Disorders; Classroom Strategies; Mental Health First Aid; and Help-Seeking), had excellent content validity with the expert inter-rater agreement Kappa of 0.932 (near-perfect agreement) (Elyamani et al., 2024).

The implications of the findings are three-fold. The findings highlight the need for randomized controlled trials and follow-up studies to provide definitive evidence on the long-term and causal impact of MHL on different Indian school settings (Liang et al., 2026; Liao et al., 2023; Prabhu et al., 2025). To ensure sustainability, embedding MHL into school culture, using a whole-school approach, mental health champions, and involving parents is a critical practice to develop in the initial stages of implementing MHL (Patel et al., 2018; O’Reilly et al., 2018; Margaretha et al., 2023). Policy, pre-service curriculum integration (D.Ed./B.Ed./M.Ed.) and integration with national initiatives like Rashtriya Kishor Swasthya Karyakram are important factors for institutionalization (Mastrorio et al., 2020; Parikh et al., 2019; Roy et al., 2019; Whitley & Gooderham, 2016).

Overall, the validated module provides teachers with the skills needed to be culturally responsive, policy-informed, and practically oriented, thereby helping schools to become resilient environments of support where adolescent well-being is protected and India’s public-health efforts are reinforced, even in low-resource settings (Aslam et al., 2025; Barry et al., 2013; Huang et al., 2022; Kahi et al., 2025; Skar et al., 2024).

## Data Availability

The author has secured the data. Deidentified individual participant data (including data dictionaries) will be made available, in addition to study protocols, the statistical analysis plan, and the informed consent form. The data will be made available upon publication to researchers who provide a methodologically sound proposal to achieve the goals of the approved proposal.

